# Post-Operative Atrial Arrhythmias after Lung Transplantation: A Single Center Analysis of Risk Factors, Management, and Outcomes

**DOI:** 10.1101/2025.07.01.25330683

**Authors:** Daniel M. Chen, Ambalavanan Arunachalam, Graham Peigh, Bradley P. Knight, Chitaru Kurihara, Mrinalini Venkata Subramani, Catherine Myers, Alan Betensley, Krishnan Warrior, Bradford C. Bemiss, Amanda Kamar, Mohamed Al-Kazaz

## Abstract

**Background:** Following lung transplant, many patients develop post-operative atrial fibrillation (AF) or atrial flutter (AFL)–collectively defined as post-operative atrial arrhythmia (POAA). While early (in-hospital) POAA is linked to increased mortality, late (post-discharge) POAA is poorly understood. Risk factors and real-world management strategies for POAA are not well characterized. We aim to evaluate POAA incidence and timing, identify risk and protective factors, and assess associations with mortality and long-term outcomes.

**Methods:** This retrospective cohort study included 233 adult lung transplant recipients at Northwestern Memorial Hospital (2014–2024), excluding those with pre-existing AF or AFL. POAA was confirmed by ECG or Holter. Adjusted logistic and Cox regressions identified predictors of POAA and mortality.

**Results:** Of 233 recipients (age 58.6±12.8y, 45.1% female), POAA occurred in 29.6% (53.6% AF, 24.6% AFL, 21.7% both). Both overall (HR4.06, 95%CI[2.21–7.46]) and late (HR3.01, 95%CI[1.39–6.53]) POAA were significantly associated with mortality. 14.5% of patients with POAA required hospital-based arrhythmia management and 66.6% underwent additional rhythm control (2.9%% DCCV, 46.4% AAD initiation, 17.4% both). Preoperative pulmonary arterial hypertension (PAH) was associated with less POAA (OR0.31, 95%CI[0.14–0.62]). Postoperative beta-blocker use was associated with 73% reduced POAA (HR0.27, 95%CI[0.09–0.80]).

**Conclusions:** This study is among the first to associate late POAA with mortality and define arrhythmia-related hospitalization rates after POAA. Postoperative beta-blockers were associated with significantly less POAA, a novel finding in lung transplant recipients. Finally, preoperative PAH’s association with less POAA might reflect reduced cardiac strain post-transplant.

## INTRODUCTION

Lung transplantation is lifesaving for many patients with end stage pulmonary disease. However, lung transplant recipients continue to experience a high rate of post-operative complications, such as primary graft dysfunction, chronic lung allograft dysfunction, and increased risk of infection^1^. POAA is one the most common complications, affecting approximately 20%^2^ to 40%^3^ of patients.

Despite its prevalence, the long-term clinical implications of POAA are poorly understood. Most studies focus on early POAA during hospitalization, with limited data on late POAA occurring after hospital discharge^4^. While early POAA has been consistently associated with increased mortality^5–9^, only one study has reported a similar association in late POAA^6^. Additionally, the risk factors for POAA in lung transplant recipients are not fully understood. Although some studies suggest associations with age^2,7–14^, COPD^12,15^, CAD^8,16,17^, male sex^8,10,13,14^, and single vs. double lung transplant^3,15,18^, findings remain inconsistent. Management of POAA in lung transplant patients is further complicated by a lack of evidence-based guidelines, with most treatment decisions informed by guidelines for other thoracic^12^ or general surgical^19^ patients. Specifically, there are limited data on the outcomes after initiation of beta blockade or oral anticoagulation (OAC) in this population. Finally, evidence is sparse regarding the association between POAA and outcomes such as stroke^2,8,16^, myocardial infarction, or hospitalization for heart failure or arrhythmia care. Contemporary studies are especially needed to reflect the evolving lung transplant population, which now includes patients with COVID-related lung disease accounting for 8.7% of all lung transplants in the United States from August 2020 to June 2022^20^.

Given the frequency and potential impact of POAA after lung transplant, it is critical that we gain a better understanding of its risk factors and clinical implications. This retrospective study aims to characterize the incidence, risk factors, outcomes, and treatment options for POAA in a large cohort. We hypothesize that POAA will be associated with significant morbidity and mortality, with an increased long-term risk of conditions like stroke, MI, and new hospitalizations for atrial arrhythmia and heart failure.

## METHODS

### Study Design and Population

We conducted a retrospective analysis of all patients enrolled in a lung transplant repository at Northwestern Memorial Hospital, which began enrolling patients in February 2021 (STU00212120). To ensure at least six months of follow up, we applied a surgical cut-off date of 10/23/2024, yielding an initial cohort of 318 patients. Patients were excluded if they did not ultimately undergo lung transplant (n = 17), lacked clinical documentation of the index hospitalization (n = 2), had a pre-existing history of atrial arrhythmia (n = 64), or underwent combined heart-lung transplant (n = 2), leaving a final analytic sample of 233 patients. Patients were followed from the time of transplant until death or the date of chart review completion (4/23/2025). This study was approved by the Northwestern Institutional Review Board (STU00223074). The requirement for informed consent was waived by the IRB. All procedures were conducted in accordance with institutional guidelines and the ethical standards of the 1964 Helsinki declaration. This manuscript was prepared in accordance with the Strengthening the Reporting of Observational Studies in Epidemiology (STROBE) guidelines.

### Definitions

POAA was defined as atrial fibrillation or flutter occurring after lung transplant, confirmed by 12-lead ECG or ambulatory rhythm monitoring and interpreted by a cardiologist. POAA was further classified as early (defined as occurring during the index hospitalization) and late (occurring after hospital discharge). COPD and alpha-1 antitrypsin deficiency were classified as obstructive lung disease. Interstitial pulmonary fibrosis, hypersensitivity pneumonitis, systemic autoimmune rheumatic disease-associated interstitial lung disease (SARD-ILD), and unspecified ILD were classified as restrictive lung disease. PAH was defined by mPAP>21 mmHg, PCWP<15, and PVR>2.

### Data Collection

Data on patient demographics, pre-transplant comorbidities, post-transplant clinical course, and post-discharge outcomes were collected through a thorough review of the medical record. For outcome analyses, POAA was only considered present if it occurred before the outcome of interest. Several steps were taken to minimize potential sources of bias. To minimize observer bias, standardized protocols were used for data collection, and data collectors were not involved in patient care. To minimize misclassification bias, POAA required confirmation with 12-lead EKG or ambulatory rhythm monitor. There were no missing data for variables included in the primary analysis. Missingness was limited to variables derived from right heart catheterizations and transthoracic echocardiograms, which were not routinely available for all patients. The specific number of missing values for each variable is listed in Table S2. These data were used exclusively in a secondary mediation analysis and handled with complete case analysis. Hemodynamic data was determined by the last pre-transplant right heart catheterization. Functional cardiac data was determined by the echocardiogram obtained when the patient was listed for lung transplant.

### Statistical Analysis

All statistical analyses were performed using R version 4.5.0. Generative AI was used to suggest appropriate statistical tests and help write code in R. To determine which factors were associated with POAA, we initially performed univariate logistic regressions, and then included the factors with a p-value of < 0.25 and those with clinical plausibility in a multivariate regression model. All outcome variables had low incidence rates, so we used Fisher’s exact analysis to compare outcomes between patients with and without POAA. Univariate and multivariate Cox logistic regressions were used to determine which variables were associated with mortality. Finally, a multivariate time-dependent Cox regression was used to determine the effect of post-operative beta blockers on POAA risk. Most averages are reported as mean ± SD, with median (IQR) reported for small or skewed samples. The p-values for hypothesis-driven analyses (including the association between POAA and mortality, the effect of post-operative beta blocker use on POAA, and outcomes associated with POAA) were not adjusted for multiple comparisons due to the small number of prespecified tests. All other statistical tests were exploratory and hypothesis generating. The corresponding author has full access to all the data in the study and takes responsibility for its integrity and the data analysis.

## RESULTS

### Descriptive Characteristics and Risk Factors for POAA

The characteristics of the 233 patients are summarized in Table 1. On average, patients with POAA were older, had more smoking pack years, were more likely to have CAD, and less likely to have hemodynamically defined PAH than patients without POAA. Indications for transplant included COVID-related lung disease (n = 19; 8.2%), 21.0% obstructive lung disease (n = 49; 21.0%), restrictive lung disease (n = 106; 45.5%), pulmonary hypertension (n = 16, 6.9%), and other (n = 42; 18.0%). Of the 69 patients that developed POAA, there were 37 (53.6%) patients that developed AF, 17 (24.6%) patients that developed AFL, and 15 (21.7%) patients that developed both. There were 18 (26.1%) patients that developed POAA (27.8% AF; 50% AFL; 22.2% both) after hospitalization, at a median time of 215.5 days after transplant (IQR 65.5 – 505.5 days). Incidence of early and late AF and AFL are summarized in Table 2.

**Table 1:**
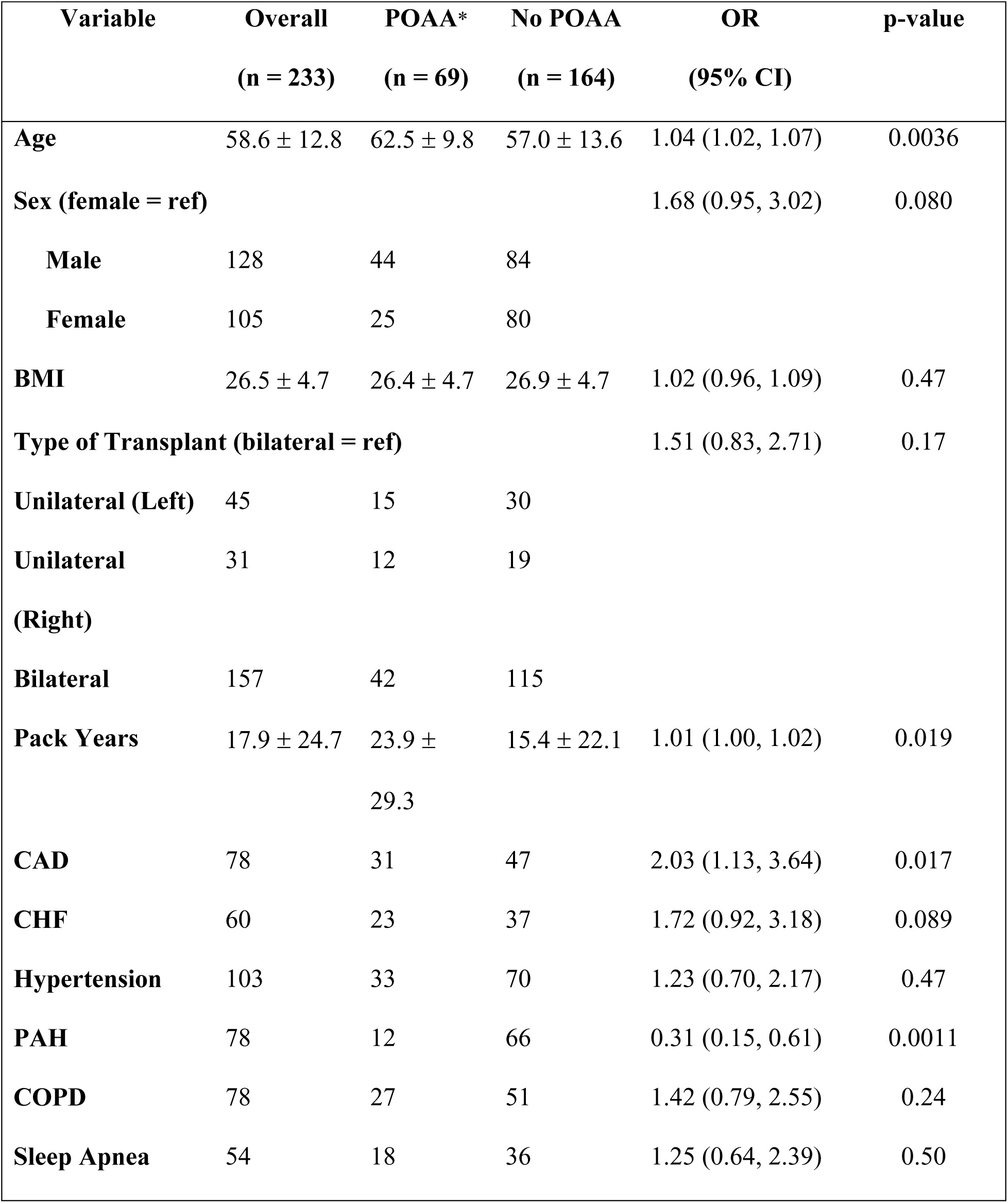
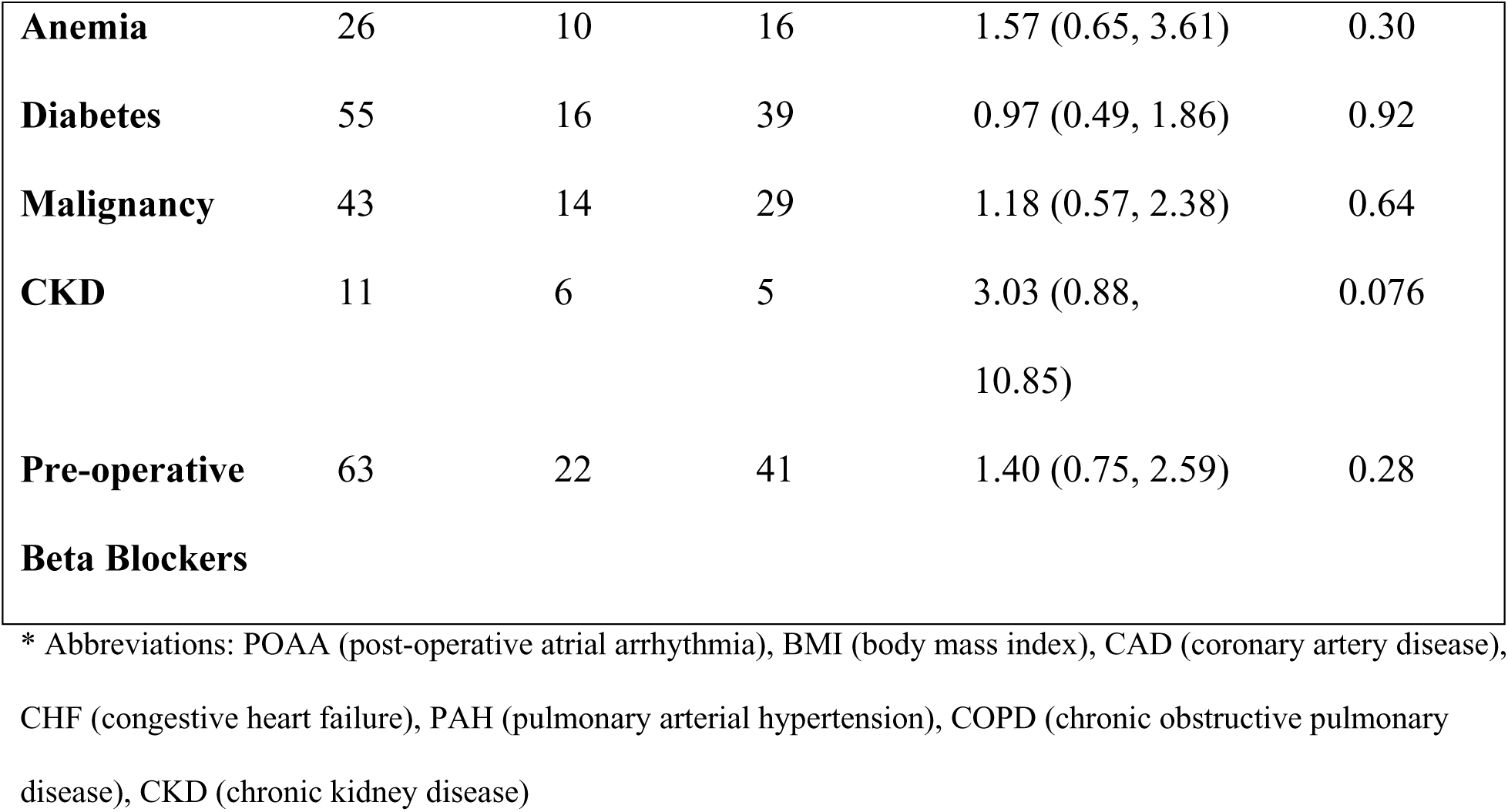
Descriptive characteristics and unadjusted risk factors for POAA.

**Table 2:** Incidence of Early and Late Post-Operative AF and AFL.

| **Arrhythmia Type** | **Total and Incidence (% of cohort, n = 233)** | **Early (During Index Hospitalization)** | **Late (After Discharge)** |
| --- | --- | --- | --- |
| **Total POAA\*** | 69 (29.6%) | 51 (21.9%) | 18 (7.7%) |
| **AF** | 37 (15.9%) | 32 (13.7%) | 5 (2.1%) |
| **AFL** | 17 (7.3%) | 8 (3.4%) | 9 (3.9%) |
| **Mixed AF/AFL** | 15 (6.4%) | 11 (4.7%) | 4 (1.7%) |

\* Abbreviations: POAA (post-operative atrial arrhythmia), AF (atrial fibrillation), AFL (atrial flutter)

Table 1 also describes which risk factors were associated with POAA in unadjusted univariate models. Age, number of pack years, and CAD were associated with more POAA, while PAH was found to have a protective association. All factors with a p-value of < 0.25 on univariate analysis were included in a multivariate regression model (Figure 1). On multivariate analysis, a history of PAH was found to have a protective association against developing POAA (OR 0.31, 95% CI 0.14 – 0.62, p = 0.0015). The other factors did not have statistically significant associations (Table S1). Because there was a protective association between PAH and POAA, we performed a nonparametric bootstrapped mediation analysis with 5,000 iterations. Several candidate mediators were assessed, including both hemodynamic (mPAP, PCWP, CO, PVR) and functional (RVSF, TR) variables. None of the mediators demonstrated a statistically significant mediation effect (Table S2), and the analyses could not be completed for the functional measures (RVSF, TR, RV size) because bootstrap iterations failed to converge.

**Figure 1:**
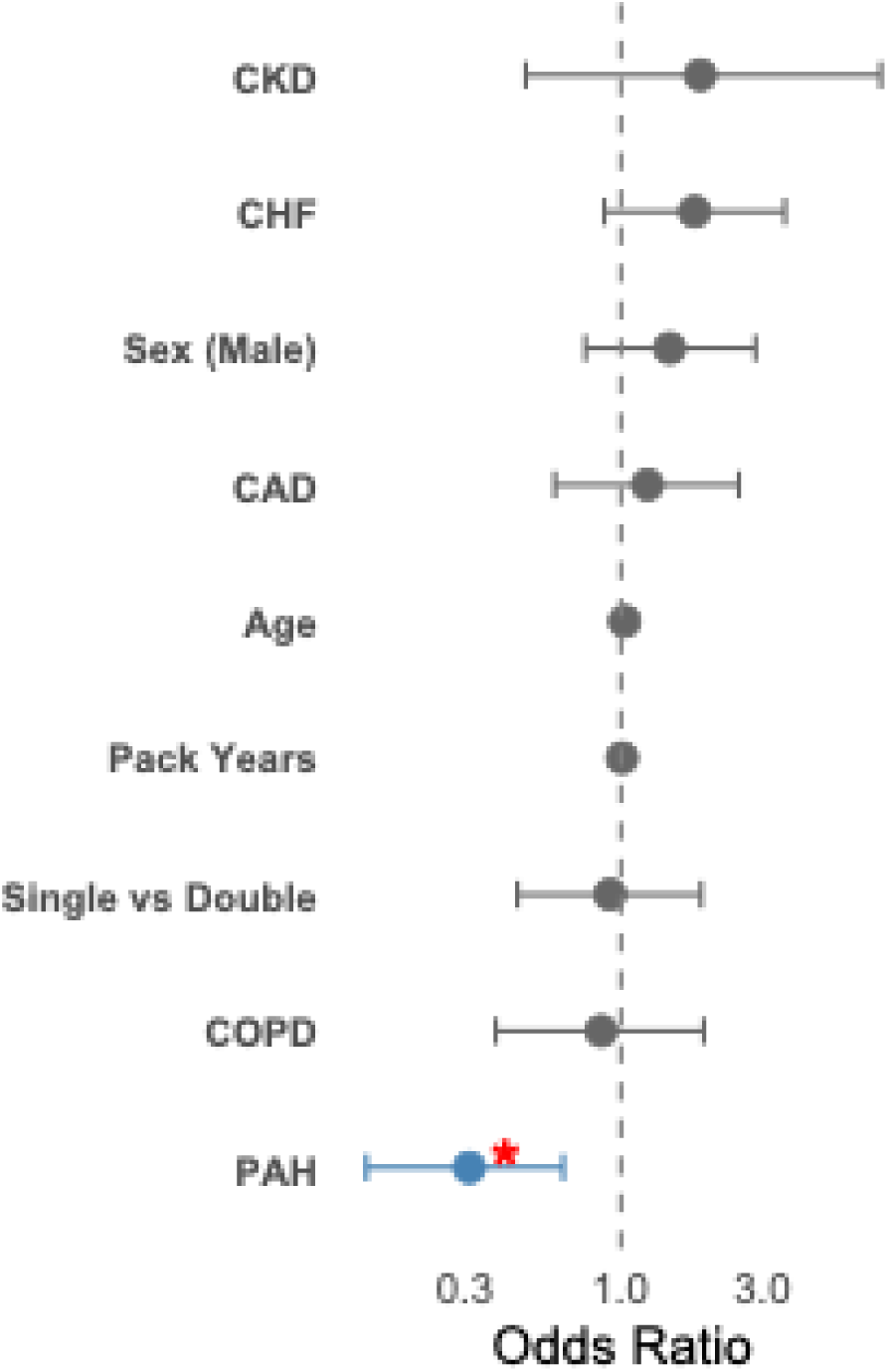
Adjusted risk factors for POAA. Multivariate regression model identifying risk factors for POAA. A preoperative history of PAH was found to have a protective association against POAA (OR 0.31, 95% CI 0.14–0.62, p = 0.0015). Abbreviations: CKD (chronic kidney disease), CHF (congestive heart failure), CAD (coronary artery disease), COPD (chronic obstructive pulmonary disease), PAH (pulmonary arterial hypertension).

We ran separate univariate and multivariate models for early and late POAA, and found that age (OR 1.04, 95% CI 1.01–1.08, p = 0.010) and PAH (OR 0.35, 95% CI 0.15–0.74, p = 0.0085) were both associated with early POAA in univariate models. There were no significant predictors of early arrhythmia in our multivariate model, and there were no significant predictors of late POAA in univariate models (Tables S3-S5). We also ran separate analyses for post-operative AF and AFL, and found that age (OR 1.05, 95% CI 1.02–1.10, p = 0.011) was associated with post-operative AF in univariate models. For AFL, PAH (OR 0.11, 95% CI 0.01–0.57, p = 0.036) was the only significant univariate predictor, and for mixed AF/AFL, recipient pack years was the only significant univariate predictor (OR 1.02, 95% CI 1.00–1.04, p = 0.038). In multivariate models, there were no significant predictors of AF, AFL or mixed AF/AFL (Tables S6-S9).

We attempted to perform a propensity match analysis to determine whether single or double lung transplant was more likely to be associated with POAA. However, a 1:2 nearest neighbor propensity match failed due to non-overlap between propensity scores. A Kaplan-Meier analysis was performed, and there was no statistically significant difference in time to atrial arrhythmia between single and double lung transplant patients (HR 1.40, 95% CI 0.87–2.28, p = 0.17, Figure S1). The median length of stay was 17 days (IQR 11–31) in patients with POAA and 15 days (IQR 11–26) in those without POAA. The difference was not statistically significant (p = 0.14, Wilcoxon rank sum test).

### Management of POAA and outcomes associated with POAA

Management strategies for the 69 patients who developed POAA are described in Table 3. Among the patients who developed POAA during hospitalization, only one patient was discharged on anticoagulation (CHA_2_DS_2_-VASC 5, no history of stroke). 33 patients were started on anticoagulation during follow up. Anticoagulation for POAA was initiated 41 (IQR 8 – 173.25) days post-transplant and was continued for an average of 17.3 ± 13.2 months. More patients were anticoagulated for DVT/PE (n = 25, 36.2%) than AF/AFL (n = 7, 10.1%). Prescription of anticoagulation by CHA_2_DS_2_-VASC score is described in Table S10.

**Table 3:** Management strategies for POAA.

| **Management Strategy** | **Overall POAA<sup>†</sup>**<br>**(n = 69)** | **Early POAA**<br>**(n = 51)** | **Late POAA**<br>**(n = 18)** |
| --- | --- | --- | --- |
| **Rate only** | 16 | 9 | 7 |
| **AAD only** | 9 | 8 | 1 |
| **Rate, AAD** | 23 | 18 | 5 |
| **Rate, DCCV** | 2 | 1 | 1 |
| **AAD, DCCV** | 3 | 3 | 0 |
| **Rate, AAD, DCCV** | 9 | 8 | 1 |
| **None** | 7 | 4 | 3 |

† Abbreviations: POAA (post-operative atrial arrhythmia), AAD (antiarrhythmic drug), DCCV (direct current cardioversion)

Table 4 compares the incidence of stroke, MI, and new hospitalization for AA or heart failure among patients that developed POAA and patients that did not develop POAA. We used Fisher’s exact test because of the low incidence rates of all outcome variables. Two patients had a stroke after developing early POAA. Both patients were not on anticoagulation at the time. One patient had a CHA_2_DS_2_-VASC score of 6, with stroke occurring 6 days after transplant and 2 days after POAA; and the other patient had a CHA_2_DS_2_-VASC of 5, with stroke occurring 71 days after transplant and 50 days after POAA. Of the patients that developed POAA, 17.4% (12/69) presented to the ED or were hospitalized for arrhythmia during follow-up, compared to 0% of patients that did not develop POAA. Of the 12 patients, 5 had early POAA and 7 had late POAA.

**Table 4:**
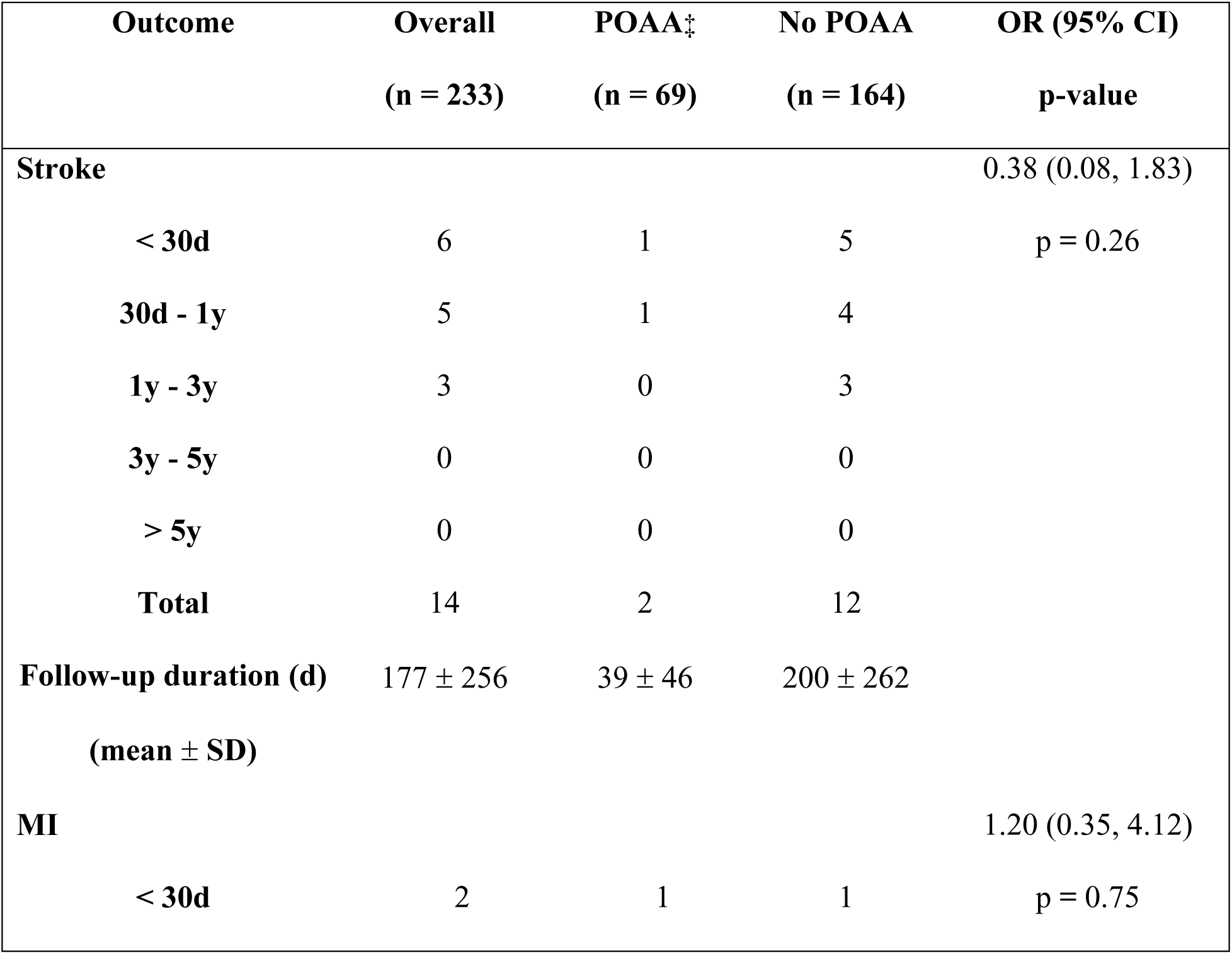
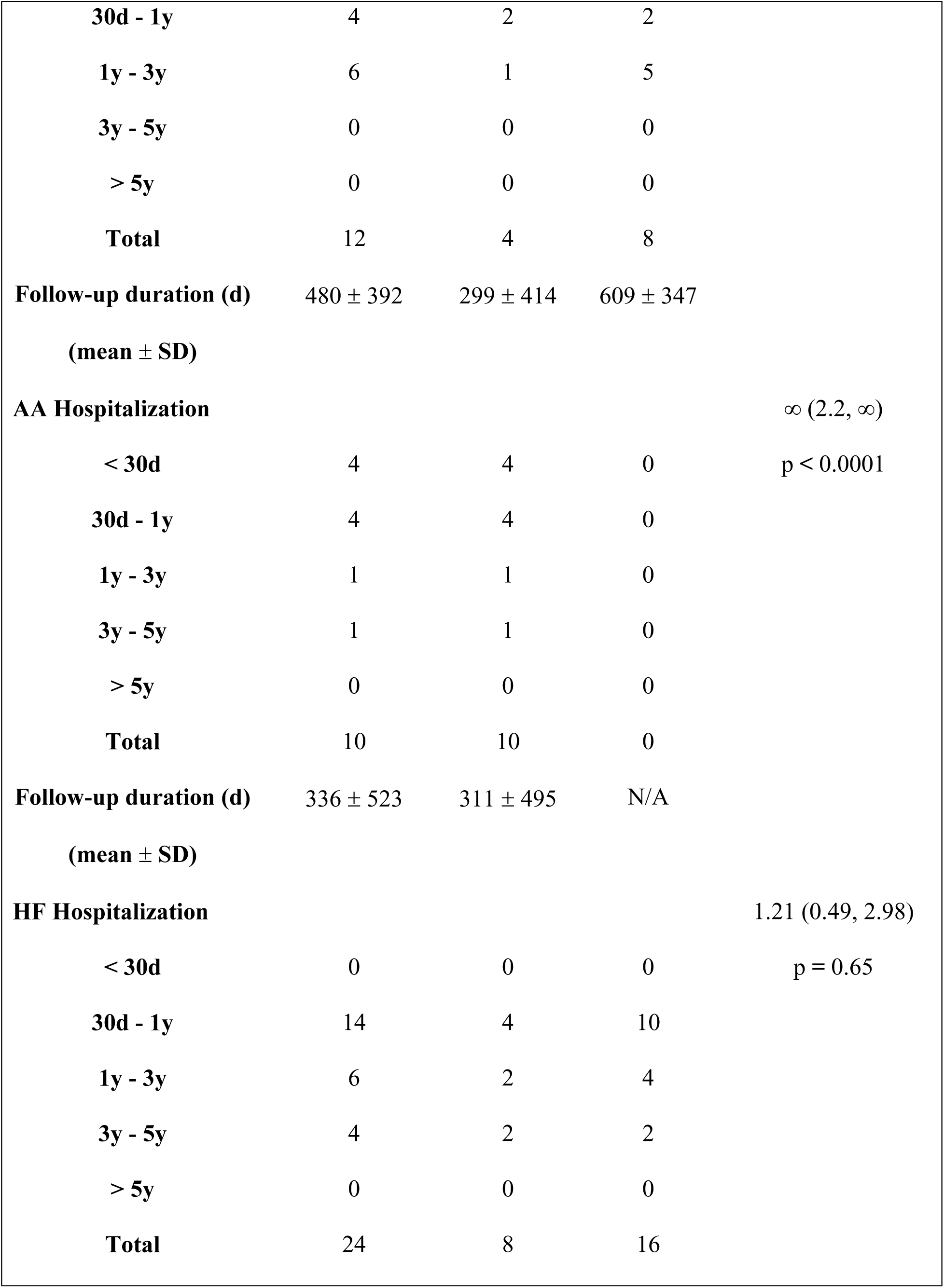
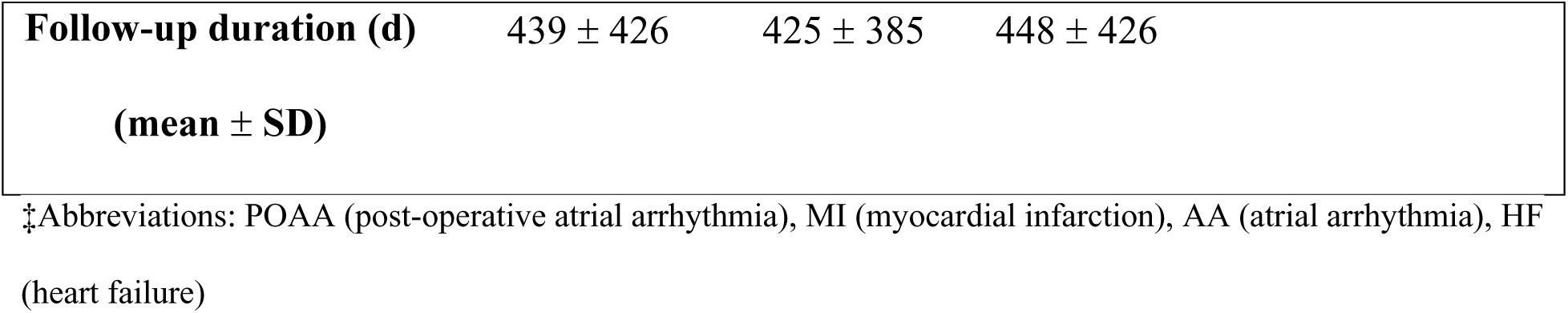
Outcomes associated with POAA.

| **Outcome** | **Overall**<br>**(n = 233)** | **POAA<sup>†</sup>**<br>**(n = 69)** | **No POAA**<br>**(n = 164)** | **OR (95% CI)**<br>**p-value** |
| --- | --- | --- | --- | --- |
| **Stroke** |  |  |  | 0.38 (0.08, 1.83) |
| < 30d | 6 | 1 | 5 | p = 0.26 |
| 30d - 1y | 5 | 1 | 4 |  |
| 1y - 3y | 3 | 0 | 3 |  |
| 3y - 5y | 0 | 0 | 0 |  |
| > 5y | 0 | 0 | 0 |  |
| **Total** | 14 | 2 | 12 |  |
| **Follow-up duration (d)** | 177 ± 256 | 39 ± 46 | 200 ± 262 |  |
| **(mean ± SD)** |  |  |  |  |
| **MI** |  |  |  | 1.20 (0.35, 4.12) |
| < 30d | 2 | 1 | 1 | p = 0.75 |
| **30d - 1y** | 4 | 2 | 2 |  |
| **1y - 3y** | 6 | 1 | 5 |  |
| **3y - 5y** | 0 | 0 | 0 |  |
| **> 5y** | 0 | 0 | 0 |  |
| **Total** | 12 | 4 | 8 |  |
| **Follow-up duration (d)** | 480 ± 392 | 299 ± 414 | 609 ± 347 |  |
| **(mean ± SD)** |  |  |  |  |
| **AA Hospitalization** | | | | ∞ (2.2, ∞) |
| **< 30d** | 4 | 4 | 0 | p < 0.0001 |
| **30d - 1y** | 4 | 4 | 0 |  |
| **1y - 3y** | 1 | 1 | 0 |  |
| **3y - 5y** | 1 | 1 | 0 |  |
| **> 5y** | 0 | 0 | 0 |  |
| **Total** | 10 | 10 | 0 |  |
| **Follow-up duration (d)** | 336 ± 523 | 311 ± 495 | N/A |  |
| **(mean ± SD)** |  |  |  |  |
| **HF Hospitalization** |  |  |  | 1.21 (0.49, 2.98) |
| **< 30d** | 0 | 0 | 0 | p = 0.65 |
| **30d - 1y** | 14 | 4 | 10 |  |
| **1y - 3y** | 6 | 2 | 4 |  |
| **3y - 5y** | 4 | 2 | 2 |  |
| **> 5y** | 0 | 0 | 0 |  |
| **Total** | 24 | 8 | 16 |  |

|  |  |  |  |
| --- | --- | --- | --- |
| **Follow-up duration (d)** | 439 ± 426 | 425 ± 385 | 448 ± 426 |
| **(mean ± SD)** |  |  |  |

‡Abbreviations: POAA (post-operative atrial arrhythmia), MI (myocardial infarction), AA (atrial arrhythmia), HF (heart failure)

### Predictors of Mortality

Table S11 describes which variables are associated with mortality in univariate Cox regression models. Single lung transplant, length of hospitalization, CKD, and POAA (overall, early, and late) were all found to be significantly associated with increased mortality. PAH had a protective association against mortality.

All factors with p < 0.25 in our unadjusted model were included in a multivariate regression analysis (Figure 2). Significant predictors of mortality in the adjusted model included single vs. double lung transplant, length of hospital stay and POAA. The multivariate regression analysis was re-run with early and late POAA. Late POAA was also found to be a significant predictor of mortality (HR 4.37, 95% CI 1.90 – 10.06, p = 0.00051). There was no significant association between early POAA and mortality (Table S12).

**Figure 2:**
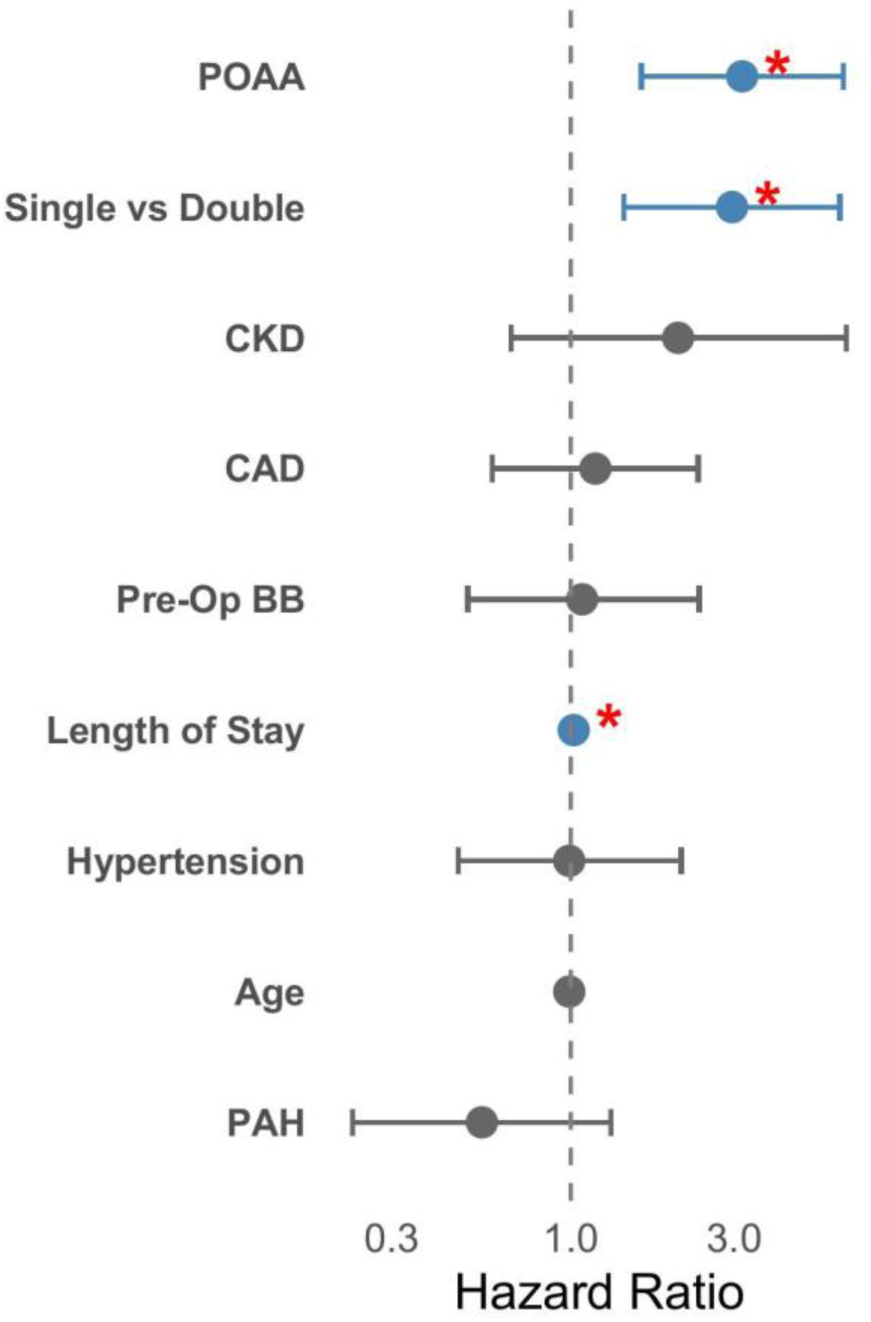
Adjusted risk factors for mortality. Multivariate regression model identifying factors associated with post-transplant mortality. The development of POAA was associated with increased mortality (HR 3.09, 95% CI 1.57–6.08, p = 0.0011), as were single lung transplant versus double (HR 2.93, 95% CI 1.41– 6.07, p = 0.0039), and longer hospital length of stay (HR 1.02, 95% CI 1.01–1.03, p = 0.0000052). Abbreviations: POAA (post-operative atrial arrhythmia), CKD (chronic kidney disease), CAD (coronary artery disease), BB (beta blocker), PAH (pulmonary arterial hypertension).

**Figure 3:**
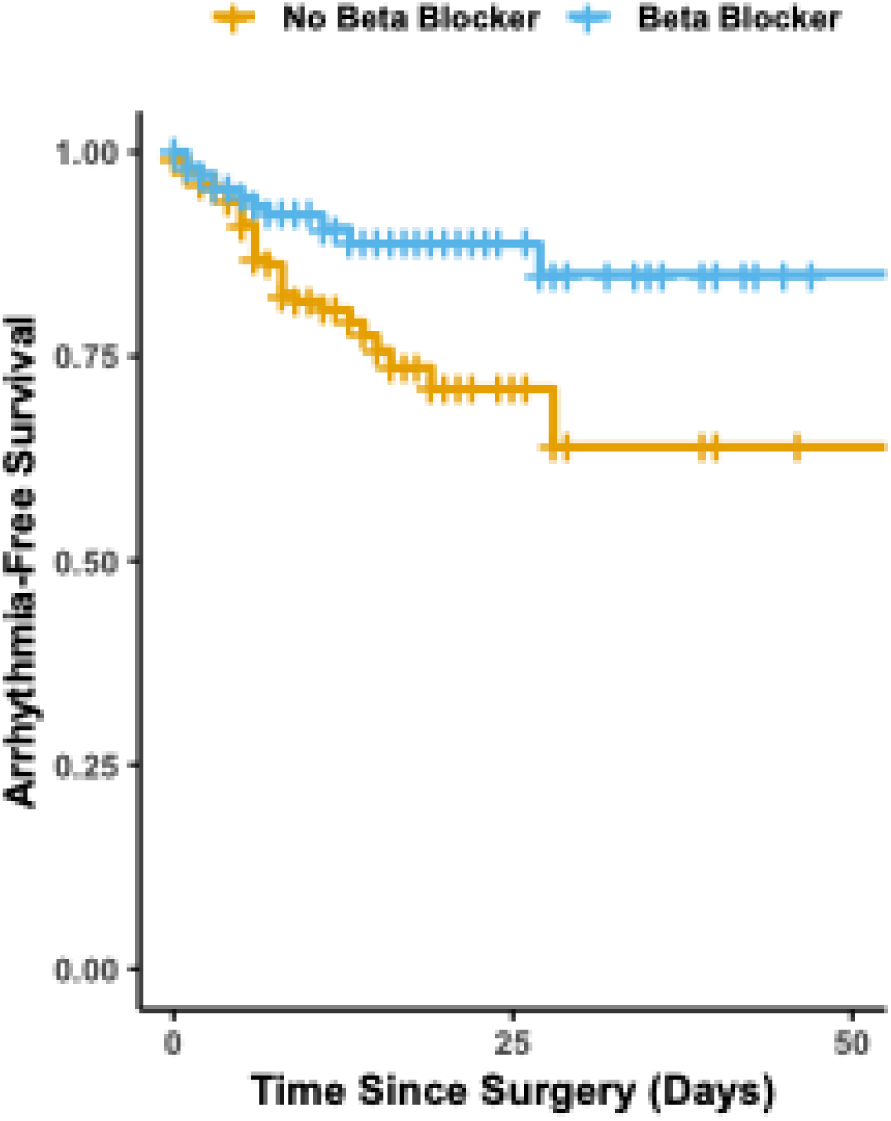
Effect of post-operative beta blockers on POAA risk. Kaplan-Meier curve stratifying arrhythmia-free survival by post-operative beta blocker use. Patients who received post-operative beta blockers demonstrated a lower incidence of arrhythmia compared to those who did not. In a time-dependent Cox proportional hazards model, post-operative beta blocker use was associated with a significantly reduced hazard of developing arrhythmia (HR 0.27, 95% CI 0.09–0.80, p = 0.018).

### Effects of post-operative beta blockers on POAA risk

A total of 141 patients received post-operative beta blockers after lung transplant surgery, including 82 who were beta-blocker naïve prior to transplant. These beta blockers were started an average of 8 ± 6 days after transplant. We ran a time-dependent Cox regression analysis to see whether post-operative beta blocker use had a protective association against POAA (Table 5). Post-operative beta blocker use was associated with a significantly reduced hazard of developing arrhythmia (HR 0.27, 95% CI 0.09 – 0.80, p = 0.018). A Kaplan-Meier survival curve is provided as a visual depiction of arrhythmia-free survival by post-operative beta blocker use (Figure S1).

**Table 5:** Adjusted effect of post-operative beta blockers on POAA risk.

| **Variable** | **HR (95% CI)** | **p-value** |
| --- | --- | --- |
| **Post-Operative Beta Blocker** | 0.27 (0.093–0.80) | 0.018 |
| **Beta Blockers Prior to Admission** | 1.32 (0.42–4.15) | 0.63 |
| **Age** | 1.05 (1.00–1.09) | 0.051 |
| **Sex (Male)** | 2.84 (1.09–7.44) | 0.033 |
| **Pack Years** | 1.00 (0.99–1.02) | 0.68 |
| **CAD§** | 0.76 (0.28–2.10) | 0.60 |
| **CHF** | 1.57 (0.53–4.65) | 0.42 |
| **HTN** | 0.73 (0.28–1.91) | 0.52 |
| **PAH** | 0.45 (0.15–1.40) | 0.17 |
| **Anemia** | 1.53 (0.46–5.10) | 0.49 |
| **CKD** | 1.16 (0.17–7.81) | 0.88 |

§ Abbreviations: CAD (coronary artery disease), CHF (congestive heart failure), HTN (hypertension), PAH (pulmonary arterial hypertension), CKD (chronic kidney disease)

## DISCUSSION

In this single-center retrospective cohort study of 233 patients, the incidence of postoperative atrial arrhythmia was 29.6%, with the majority of arrhythmia being AF. Post-operative beta blockers and a history of pre-operative PAH were associated with less POAA in adjusted models. Overall and late (but not early) POAA were both found to be associated with increased mortality in an adjusted model, and 17.4% of patients with POAA presented to the ED or were hospitalized for arrhythmia during follow-up. Management strategies for POAA tended to favor rate control, and only 7/69 patients with POAA were prescribed anticoagulation during follow-up. There were two POAA patients not on anticoagulation who suffered an ischemic stroke.

In all surgical patients, there are several arrhythmogenic mechanisms, including increased sympathetic drive, fluid shifts, and electrolyte disturbances^21^. In lung transplantation, there are a unique set of considerations, most notably the removal of the native pulmonary veins and the anastomosis of donor pulmonary veins to the left atrium^10^. Although POAA has generally been considered a self-limiting and benign phenomenon, our study is part of a growing body of evidence that challenges this view. In our study, 7.7% of our patients developed POAA after hospitalization at a median time of 215.5 days after transplant (IQR 65.5 – 505.5 days).

Additionally, our study is one of only two^5^ to report an association between late POAA and increased mortality. We acknowledge that atrial arrhythmias may not have a causal effect on mortality and that there may be several confounders. Nevertheless, these findings suggest a possible role for long-term rhythm monitoring in this population to better characterize arrhythmia burden and associated risks. Because patients are not routinely monitored for arrhythmia after lung transplant, it is likely that our study did not capture cases of subclinical arrhythmia, and more data is needed to assess true arrhythmia burden and clinical significance.

Our study is the first to quantify arrhythmia-related emergency department visits and hospitalizations in this population, revealing that 17.4% of patients with POAA experienced such events. Future studies should be designed to evaluate longer-term outcomes such as myocardial infarction, heart failure hospitalizations, and cerebrovascular accidents, which our study attempted to investigate but was underpowered to assess.

Notably, post-operative beta blocker therapy had a significant protective association against early (in-hospital) POAA. We recognize that patients who received beta blockers were likely hemodynamically stable enough to tolerate them, which may introduce selection bias.

Nonetheless, to our knowledge, this is the first study to demonstrate a protective association of beta blockers specifically in lung transplant patients. While another study reports no significant difference in POAA between patients who were or were not taking beta blockers at time of transplant^7^, their analysis uses an unadjusted Chi-square test, and we believe that this study’s adjusted time-dependent Cox regression better accounts for covariates and temporal dependence.

The management of POAA in lung transplant recipients remains challenging due to a lack of transplant-specific guidelines and conflicting evidence from broader surgical populations. While the use of prophylactic beta blockers is well studied in cardiac surgery patients^22,23^, and endorsed by the 2020 ESC guidelines (Class 1a)^24^, their use in thoracic surgery patients remains controversial. The most recent AATS^25^ and ACC/AHA guidelines discourage the use of beta-blockers in beta-blocker naïve patients, given the increased stroke and mortality risk demonstrated in the POISE trial^26^. However, this trial enrolled all non-cardiac surgery patients, and in thoracic surgery populations, three RCTs^27–29^ and two more recent large meta-analyses^30,31^ demonstrate that low-dose beta-blockers are effective and well tolerated. These findings have not translated to updated recommendations. We argue that data from general non-cardiac surgery patients cannot be extrapolated to thoracic surgery patients, who experience a substantially higher incidence of POAA. The role of amiodarone is also unclear, with some studies reporting that it increases the risk of pulmonary toxicity^32^ and mortality^8,33^, while other studies advocating for its use as a first-line agent to treat POAA after lung transplant^12^. Starting anticoagulation is also challenging, given post-operative bleeding risk and the need for frequent surveillance bronchoscopies, but potentially beneficial, given that two patients with POAA had strokes while off anticoagulation. Overall, more targeted research is needed to guide the management of POAA in lung transplant patients. As previously mentioned, current practice relies heavily on data from general thoracic surgery, which in turn is often extrapolated from general non-cardiac surgical populations. Our findings underscore the need for lung transplant-specific studies to inform evidence-based guidelines in this high-risk group.

Finally, it was surprising that a pre-operative history of PAH had a negative association with POAA, even despite correcting for age. We suspect that for patients with PAH, lung transplant relieves pressure on the right atrium and ventricle, leading to cardiac remodeling and less susceptibility to POAA. This interpretation is supported by several studies that demonstrate reverse RV remodeling following lung transplant in this population^34–36^. Our finding is similar to that of Chaikriangkrai et al^14^, who found that mPAP was inversely associated with POAA. More data is needed to confirm this finding and determine how it may affect the management of lung transplant patients with PAH.

This study has several limitations. First, it is a retrospective, single-center study, which may limit generalizability and introduce bias related to documentation and clinical practice patterns. Second, subclinical atrial arrhythmias were likely underreported, particularly following hospital discharge, as routine rhythm monitoring was not performed. Third, the study was underpowered to detect associations with less frequent clinical outcomes such as heart failure hospitalization, stroke, or mortality. Lastly, the management of atrial arrhythmias was not standardized and was subject to variability based on physician judgment and individual patient factors, which may have influenced outcomes.

In conclusion, in this retrospective study of lung transplant patients, POAA was common, and was associated with higher mortality and increased arrhythmia-related ED visits and hospitalizations. Our novel findings challenge the traditional view of POAA as a benign and self-limited condition, particularly given the late onset of arrhythmia in some patients and its association with adverse outcomes. Postoperative beta blocker therapy was associated with a lower incidence of in-hospital POAA, which is also a novel observation in this population. Future studies should aim to assess the true arrhythmia burden in this population, evaluate long-term outcomes, and utilize standardized treatment protocols to better understand POAA management strategies.

## Data Availability

The data underlying this article will be shared on reasonable request to the corresponding author.

POAA: post-operative atrial arrhythmia
PAH: pulmonary arterial hypertension
AF: atrial fibrillation
AFL: atrial flutter
ILD: interstitial lung disease

## ACKNOWLEDGEMENTS

Generative AI was used for language refinement, with all outputs verified. We thank Ms. Mary Carns for managing the REDCap lung transplant biorepository.

## SOURCES OF FUNDING

None.

## DISCLOSURES

Dr. Graham Peigh serves as a consultant for Philips and Medtronic.

## Notes

### Funding Statement

The authors received no funding for any aspect of the submitted work.

### Author Declarations

This study was approved by the Northwestern Institutional Review Board (STU00223074). We also used a lung transplant repository approved by the same Institutional Review Board (STU00212120).

